# Personalized azithromycin treatment rules for children with watery diarrhea using machine learning

**DOI:** 10.1101/2024.10.27.24316217

**Authors:** Sara S Kim, Allison Codi, James A. Platts-Mills, Patricia Pavlinac, Karim Manji, Chris Sudfeld, Christopher P. Duggan, Queen Dube, Naor Bar-Zeev, Karen Kotloff, Samba O Sow, Sunil Sazawal, Benson O Singa, Judd Walson, Farah Qamar, Tahmeed Ahmed, Ayesha De Costa, David Benkeser, Elizabeth T Rogawski McQuade

**Affiliations:** Department of Epidemiology, Rollins School of Public Health, Emory University, Atlanta, GA, USA; Department of Biostatistics, Rollins School of Public Health, Emory University, Atlanta, GA, USA; Division of Infectious Diseases and International Health, Department of Medicine, University of Virginia, Charlottesville, VA, USA; Department of Global Health, University of Washington, Seattle, WA, USA; Department of Epidemiology, University of Washington, Seattle, WA, USA; Department of Pediatrics and Child Health, Muhimbili University of Health and Allied Sciences, Dar es Salaam, Tanzania; Department of Global Health and Population, Harvard T.H. Chan School of Public Health, Boston, MA, USA; Division of Gastroenterology, Hepatology and Nutrition, Boston Children’s Hospital, Boston, MA, USA; Department of Pediatrics, Queen Elizabeth Central Hospital, Blantyre, Malawi; International Vaccine Access Center, Johns Hopkins Bloomberg School of Public Health, Baltimore, ML, USA; Department of Pediatrics, Center for Vaccine Development and Global Health, University of Maryland School of Medicine, Baltimore; Department of Medicine, Center for Vaccine Development and Global Health, University of Maryland School of Medicine, Baltimore, ML, USA; Centre pour le Développement des Vaccins, Bamako, Mali; Center for Public Health Kinetics, New Delhi, Delhi, India; Childhood Acute Illness and Nutrition Network, Nairobi, Kenya; Kenya Medical Research Institute, Nairobi, Kenya; Departments of International Heath, Medicine and Pediatrics, Johns Hopkins University, Baltimore, MD, USA; Department of Pediatrics and Child Heath, Aga Khan University, Karachi, Pakistan; Nutrition and Clinical Services Division, International Centre for Diarrhoeal Disease Research, Dhaka, Bangladesh; Department of Maternal, Child, and Adolescent Health and Aging, World Health Organization, Geneva, Switzerland

## Abstract

**Introduction:** We used machine learning to identify novel strategies to target azithromycin to the children with watery diarrhea who are most likely to benefit.

**Methods:** Using data from a randomized trial of azithromycin for watery diarrhea, we developed personalized treatment rules given sets of diagnostic, child, and clinical characteristics, employing a robust ensemble machine learning-based procedure. For each rule, we estimated the proportion treated under the rule and the average benefits of treatment.

**Results:** Among 6,692 children, treatment was recommended on average for approximately one third of children. The risk of diarrhea on day 3 was 10.1% lower (95% CI: 5.4, 14.9) with azithromycin compared to placebo among children recommended for treatment. For day 90 re-hospitalization and death, risk was 2.4% lower (95% CI: 0.6, 4.1) with azithromycin compared to placebo among those recommended for treatment. While pathogen diagnostics were strong determinants of azithromycin effects on diarrhea duration, host characteristics were more relevant for predicting benefits for re-hospitalization or death.

**Conclusion:** The ability of host characteristics to predict which children benefit from azithromycin with respect to the most severe outcomes suggests appropriate targeting of antibiotic treatment among children with watery diarrhea may be possible without access to pathogen diagnostics.

## Background

Diarrhea is a leading cause of death globally, especially for children aged less than 5 years (1). Approximately 25% of diarrheal episodes are likely attributed to bacterial pathogens (2), which disproportionately contribute to severe outcomes including growth faltering and death (3–5). Antibiotic therapy could prevent poor outcomes (4,6,7), but in the absence of point-of-care diagnostics in most clinical facilities in low-resource settings, it is difficult to identify which episodes are bacterial and therefore could benefit from treatment. Currently, the World Health Organization (WHO) takes a syndromic approach by recommending antibiotics only for bloody diarrhea (8). However, this approach misses watery bacterial diarrheal episodes that would likely be responsive to antibiotic therapy and inadvertently results in the widespread overuse of antibiotics because the guidance is perceived as insufficient in practice (9). Strategies to better target antibiotic treatment are needed to optimize clinical outcomes and limit antibiotic resistance.

The factors that determine antibiotic treatment response may be more complex than bacterial etiology and antibiotic susceptibility. The AntiBiotics for Children with severe Diarrhea (ABCD) randomized clinical trial investigated the effects of azithromycin treatment among dehydrated or undernourished children with acute watery diarrhea (10). The trial found no overall survival benefit, but a small improvement in linear growth among children who received azithromycin. Children with diarrhea attributed to bacterial etiologies were found to have reduced risk of diarrhea on day 3 and of hospitalization or death by day 90 (6). Less expectedly, there was also reduced risk of these outcomes (albeit of smaller magnitude) among diarrheal episodes in which bacteria were detected but did not meet the quantity thresholds to be attributed to bacteria. Furthermore, bacterial etiology was not a strong determinant of the effect of azithromycin on linear growth outcomes; the effect of azithromycin was similar regardless of etiology. Together, these results suggest that the determinants of which children benefit from azithromycin treatment are complex and incompletely understood.

Other factors aside from diarrhea etiology have been related to azithromycin treatment benefit. The *Macrolides Oraux pour Réduire les Décès avec un Oeil sur la Résistance* (MORDOR) randomized trial identified heterogeneity in azithromycin effects on mortality by age and geographic setting (11). The largest effects in MORDOR were observed in the lowest resourced setting (Niger) and among the youngest age group (1-5 months), suggesting child and sociodemographic characteristics may be important determinants of the benefit of azithromycin. However, the AVENIR trial found that all children 1-59 months should be treated to reduce mortality in the youngest infant age groups (12). The biological mechanisms driving these differential treatment benefits are incompletely understood. Independent of diarrhea etiology, azithromycin is anti-inflammatory (13), which may result in non-specific benefits for malnourished or otherwise vulnerable children. Relatedly, antibiotics are indicated without regard to specific pathogens in some cases; the WHO recommends routine treatment of children with severe acute malnutrition (SAM) with a broad-spectrum antibiotic (14), and antibiotics have been considered for moderate malnutrition and environmental enteropathy (10). Although heterogeneity in azithromycin treatment effects by individual-level characteristics has been observed in prior studies, it is unknown whether and to what extent these factors are informative for targeting treatment of diarrhea in the absence of, or in addition to, diagnostic testing. If clinical and sociodemographic characteristics can sufficiently capture heterogeneity in the azithromycin treatment response, appropriate targeting of antibiotic treatment may be achievable even when point-of-care diagnostics are unavailable.

In this secondary analysis of the ABCD trial, we used a machine learning-based framework to develop and evaluate personalized treatment rules for the decision to treat watery diarrhea with azithromycin. By characterizing which children with watery diarrhea benefit from azithromycin and quantifying the expected benefits of personalized treatment rules for diarrhea, we identify novel strategies to target azithromycin to those who benefit most and to limit antibiotic overuse and resistance.

## Results

Among 6,699 children in the ABCD trial who underwent quantitative polymerase chain reaction (qPCR) testing, we included 6,692 (99.9%) children in this analysis. We excluded seven children due to missing or invalid results for bacterial pathogens to maintain consistency with a prior ABCD analysis (6). The 1,857 (27.7%) children that were missing the day 3 diarrhea outcome and 269 (4.0%) children that were missing the change in linear growth (length-for-age z-score [LAZ]) outcome were retained in the analysis. Overall, 679 children had diarrhea on day 3, and 298 children were re-hospitalized or died by day 90, of which 26 (9%) children died.

### i. Day 3 Diarrhea Outcome

For the day 3 diarrhea outcome, an average of 27.8% (95% CI: 26.7%, 28.9%) of children were recommended for treatment under the personalized treatment rule based on all known diagnostic, child, and clinical characteristics (i.e., the comprehensive rule) using a threshold of a 7% reduction in risk of day 3 diarrhea (Figure 1A; Table S4). Among those recommended for treatment, azithromycin was associated with an average 10.1% lower risk of diarrhea on day 3 (risk difference (RD), -0.101 [95% CI: -0.149, -0.054]). The effect of azithromycin was smaller among those for whom treatment was not recommended (average RD: -0.037 [95% CI: -0.063, -0.011]). The etiologies of diarrhea were different between children recommended for treatment under the comprehensive rule compared to those not recommended for treatment (Table 1). Children recommended for treatment were more likely to have diarrhea attributed to bacteria. Child-level factors and characteristics of the clinical illness course were also different depending on the treatment recommendation. Children recommended for treatment were approximately 1 month older, were more malnourished (i.e., lower length-for-age, weight-for-age, and weight-for-length z-scores), had higher average numbers of loose and solid stools in the past 24 hours, had longer illness duration, and were less likely to be dehydrated or experience vomiting compared to those not recommended for treatment (Table 1). The relationships between child-level expected benefit and select clinical and child characteristics are also shown in Figure S1.

**Figure 1:**
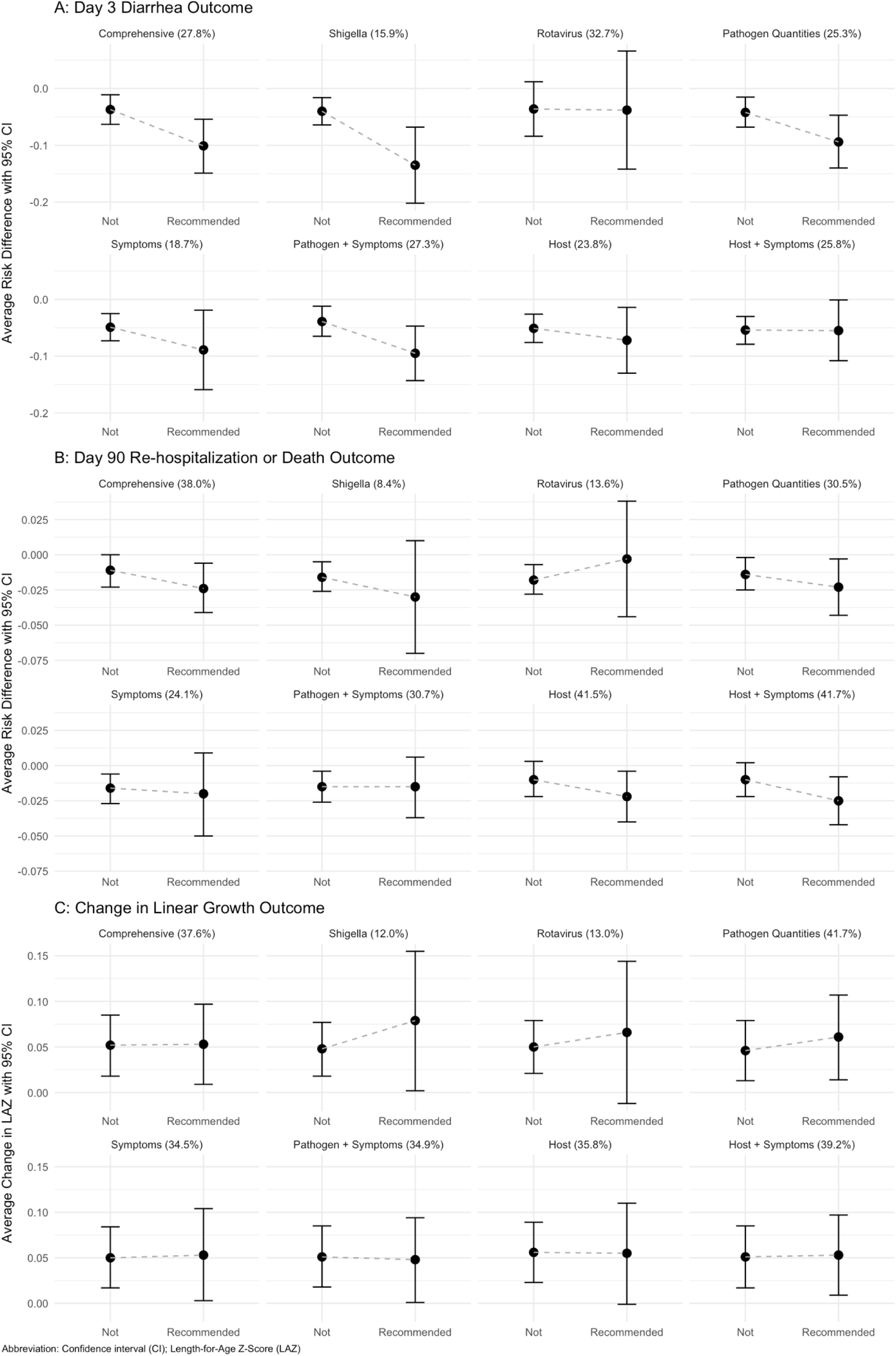
Comparison of average treatment benefit among those recommended for treatment and not recommended for treatment under each rule. The proportion recommended treatment is denoted in parentheses after the label for each rule.

**Table 1:** Descriptive characteristics by treatment recommendation under the comprehensive rule with respect to each outcome (day 3 diarrhea, day 90 re-hospitalization or death, and change in length-for-age z-score).

|  | Day 3 Diarrhea |  | Day 90 Re-Hospitalization/Death |  | Change in Linear Growth |  |
| --- | --- | --- | --- | --- | --- | --- |
|  | Not Recommended (75%) | Recommended Treatment (25%) | Not Recommended (64%) | Recommended Treatment (36%) | Not Recommended (66%) | Recommended Treatment (34%) |
|  | N (Col %); Mean (SD) | N (Col %); Mean (SD) | N (Col %); Mean (SD) | N (Col %); Mean (SD) | N (Col %); Mean (SD) | N (Col %); Mean (SD) |
| **Total** | 5,002 | 1,690 | 4,253 | 2,439 | 4,423 | 2,269 |
| **Bacteria Attributed Diarrhea** |  |  |  |  |  |  |
| Likely | 916 (18%) | 978 (58%) | 994 (23%) | 900 (37%) | 1,138 (26%) | 756 (33%) |
| Possible | 833 (17%) | 320 (19%) | 787 (19%) | 366 (15%) | 794 (18%) | 359 (16%) |
| Unlikely | 3,253 (65%) | 392 (23%) | 2,472 (58%) | 1,173 (48%) | 2,491 (56%) | 1,154 (51%) |
| **Likely *Shigella* Diarrhea** | 185 (4%) | 660 (39%) | 345 (8%) | 500 (21%) | 406 (9%) | 439 (19%) |
| **Likely ST ETEC Diarrhea** | 534 (11%) | 355 (21%) | 521 (12%) | 368 (15%) | 553 (13%) | 336 (15%) |
| **Likely tEPEC Diarrhea** | 131 (3%) | 84 (5%) | 112 (3%) | 103 (4.2%) | 161 (4%) | 54 (2%) |
| **Likely *V. Cholerae* Diarrhea** | 92 (2%) | 82 (5%) | 97 (2.3%) | 77 (3.2%) | 137 (3%) | 37 (2%) |
| **Likely *Campylobacter* Diarrhea** | 5 (<0.1%) | 1 (<0.1%) | 4 (<0.1%) | 2 (<0.1%) | 6 (0.1%) | 0 (0%) |
| **Likely *Salmonella* Diarrhea** | 38 (1%) | 14 (1%) | 26 (1%) | 26 (1%) | 24 (1%) | 28 (1%) |
| **Likely Rotavirus Diarrhea** | 1,315 (26%) | 96 (6%) | 674 (16%) | 737 (30%) | 756 (17%) | 655 (29%) |
| **Likely *Cryptosporidium* Diarrhea** | 426 (9%) | 213 (13%) | 398 (9%) | 241 (10%) | 464 (10%) | 175 (8%) |
| **Age (Months)** | 11.33 (5.11) | 12.54 (5.62) | 11.60 (5.55) | 11.69 (4.73) | 11.83 (5.42) | 11.26 (4.94) |
| **Female Sex** | 2,282 (46%) | 806 (48%) | 1,937 (46%) | 1,151 (47%) | 2,076 (47%) | 1,012 (45%) |
| **Study Site** |  |  |  |  |  |  |
| Bangladesh | 751 (15%) | 247 (15%) | 376 (8.8%) | 622 (26%) | 564 (13%) | 434 (19%) |
| India | 621 (12%) | 377 (22%) | 608 (14%) | 390 (16%) | 679 (15%) | 319 (14%) |
| Kenya | 910 (18%) | 104 (6.2%) | 855 (20%) | 159 (6.5%) | 723 (16%) | 291 (13%) |
| Malawi | 551 (11%) | 140 (8.3%) | 462 (11%) | 229 (9.4%) | 424 (9.6%) | 267 (12%) |
| Mali | 751 (15%) | 249 (15%) | 632 (15%) | 368 (15%) | 746 (17%) | 254 (11%) |
| Pakistan | 547 (11%) | 448 (27%) | 498 (12%) | 497 (20%) | 524 (12%) | 471 (21%) |
| Tanzania | 871 (17%) | 125 (7.4%) | 822 (19%) | 174 (7.1%) | 763 (17%) | 233 (10%) |
| **SES Quintile** |  |  |  |  |  |  |
| First | 658 (13%) | 206 (12%) | 223 (5.2%) | 641 (26%) | 647 (15%) | 217 (9.6%) |
| Second | 749 (15%) | 343 (20%) | 798 (19%) | 294 (12%) | 342 (7.7%) | 750 (33%) |
| Third | 710 (14%) | 361 (21%) | 586 (14%) | 485 (20%) | 658 (15%) | 413 (18%) |
| Fourth | 1,417 (28%) | 362 (21%) | 972 (23%) | 807 (33%) | 1,374 (31%) | 405 (18%) |
| Fifth | 1,468 (29%) | 418 (25%) | 1,674 (39%) | 212 (8.7%) | 1,402 (32%) | 484 (21%) |
| **# <5 Years in Household** | 1.67 (1.00) | 1.86 (1.19) | 1.63 (0.92) | 1.86 (1.24) | 1.65 (0.95) | 1.84 (1.23) |
| **Length for Age Z-Score** | -1.34 (1.33) | -1.84 (1.36) | -1.33 (1.37) | -1.71 (1.29) | -1.45 (1.33) | -1.50 (1.40) |
| **Weight for Age Z-Score** | -1.54 (1.27) | -2.03 (1.13) | -1.44 (1.32) | -2.06 (1.01) | -1.61 (1.24) | -1.76 (1.27) |
| **Weight for Length Z-Score** | -1.08 (1.26) | -1.35 (1.12) | -0.91 (1.27) | -1.55 (1.02) | -1.08 (1.22) | -1.27 (1.23) |
| **Middle-Upper Arm Circumference** | 13.23 (1.17) | 12.70 (1.13) | 13.34 (1.24) | 12.66 (0.92) | 13.18 (1.23) | 12.93 (1.07) |
| **# Loose Stools in 24 Hours** | 6.95 (3.54) | 8.31 (4.52) | 6.69 (3.33) | 8.35 (4.43) | 7.00 (3.66) | 7.88 (4.14) |
| **# Solid Stools in 24 hours** | 0.03 (0.22) | 0.16 (0.54) | 0.06 (0.032) | 0.07 (0.36) | 0.020 (0.17) | 0.015 (0.51) |
| **Illness Duration (Days)** | 2.46 (1.92) | 2.67 (2.13) | 2.46 (1.95) | 2.60 (2.02) | 2.75 (2.11) | 2.04 (1.59) |
| **Dehydration Status** |  |  |  |  |  |  |
| None | 2,063 (41%) | 990 (59%) | 1,600 (38%) | 1,453 (60%) | 1,980 (45%) | 1,073 (47%) |
| Some | 2,788 (56%) | 497 (29%) | 2,398 (56%) | 887 (36%) | 2,198 (50%) | 1,087 (48%) |
| Severe | 151 (3.0%) | 203 (12%) | 255 (6.0%) | 99 (4.1%) | 245 (5.5%) | 109 (4.8%) |
| **Vomit during Illness** | 155 (3.1%) | 30 (1.8%) | 40 (0.9%) | 145 (5.9%) | 166 (3.8%) | 19 (0.8%) |

Overall, among rules based on subsets of clinical, diagnostic, and host characteristics, rules that incorporated information on bacterial pathogen quantities most closely approximated the benefit for day 3 diarrhea observed under the comprehensive rule. For example, the average benefit among children recommended treatment by the pathogen quantities rule was a 9.4% (95% CI: 14.0%, 4.7%) average absolute reduction in risk of day 3 diarrhea and by the pathogen + symptoms rule was a 9.5% (95% CI: 14.3%, 4.7%) average absolute risk reduction (Table S4). A similar proportion of children were treated by these rules and the comprehensive rule. While the *Shigella* rule recommended treatment for a smaller proportion of children (15.9% [95% CI: 15.0%, 16.8%]), the average treatment benefit among those recommend treatment under the *Shigella* rule was larger (RD, -0.135 [95% CI: -0.202, -0.068]) than that under the comprehensive rule. Approximately 94% of children with *Shigella*-attributed diarrhea were treated, and the *Shigella* rule was more discriminating between children in that it produced the largest difference in average treatment benefits for day 3 diarrhea between those recommended for treatment and those not recommended for treatment (difference in RD: -0.095 [95% CI: - 0.166, -0.024]) (Figure 1A, Table S4). The overall average benefit among all children when treated or not treated according to the rules compared to no child treated were small and similar across rules (ranging from 1.2% to 2.6% absolute risk reduction), though the overall benefit of rules including bacterial pathogen quantities tended to be largest (Table S4). Compared to rules that did not include bacterial pathogen quantities, rules that included them generally had higher average sensitivity and specificity for identifying who should be recommended for treatment by the comprehensive (Table 2, Table S5). Finally, the child-level expected benefit under the pathogen quantities rule (concordance correlation coefficient (CCC): 0.76 [95% CI: 0.75, 0.77]) and pathogen + symptoms rule (CCC: 0.82 [95% CI: 0.81, 0.83]) were best correlated with the expected benefit under the comprehensive rule (Figure 2A). The concordance of treatment assignment between rules in relation to the child-level expected benefit and select clinical and child characteristics is shown in Figure S2.

**Figure 2:**
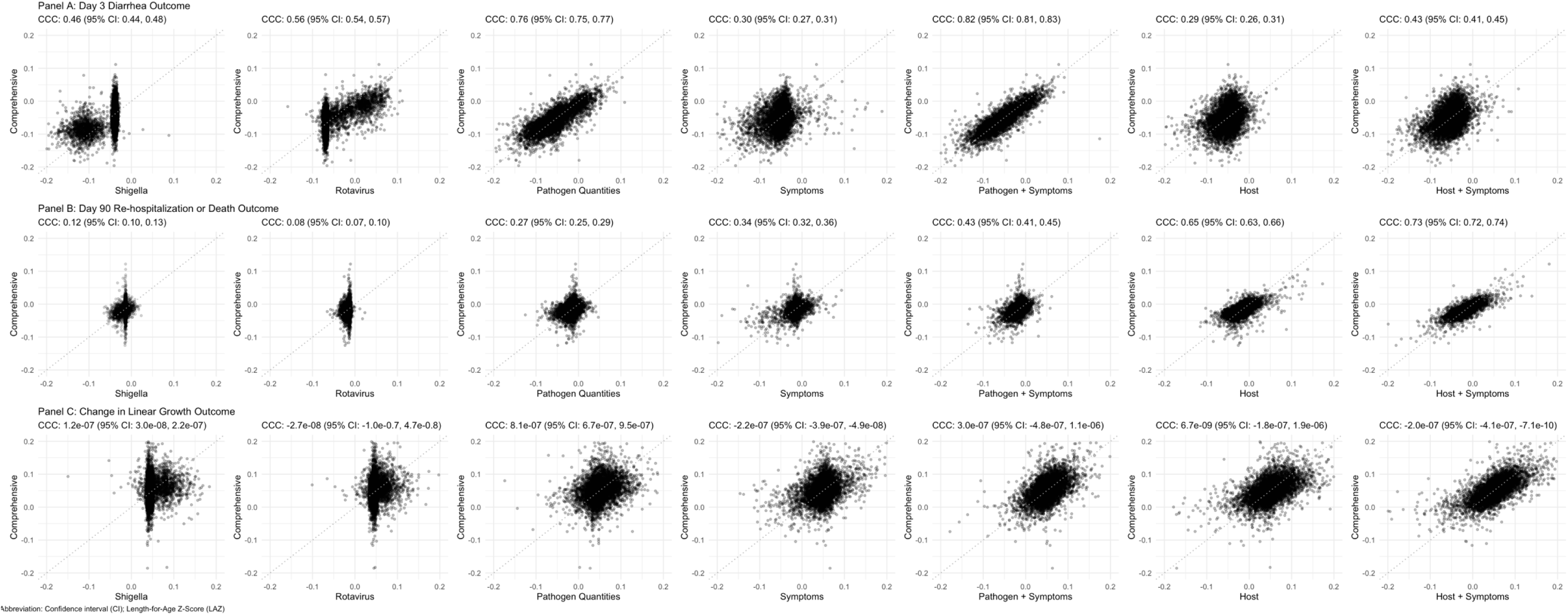
Concordant correlation coefficient (CCC) between child-level expected benefits from the comprehensive and alternative treatment rules.

**Table 2:**
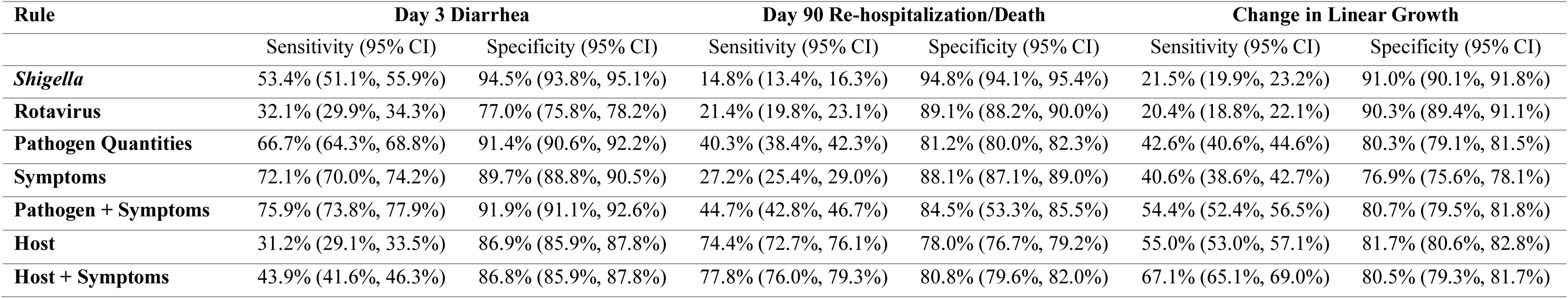
Sensitivity and specificity of each rule to identify children who were recommended treatment under the comprehensive rule.

| Rule | Day 3 Diarrhea |  | Day 90 Re-hospitalization/Death |  | Change in Linear Growth |  |
| --- | --- | --- | --- | --- | --- | --- |
|  | Sensitivity (95% CI) | Specificity (95% CI) | Sensitivity (95% CI) | Specificity (95% CI) | Sensitivity (95% CI) | Specificity (95% CI) |
| *Shigella* | 53.4% (51.1%, 55.9%) | 94.5% (93.8%, 95.1%) | 14.8% (13.4%, 16.3%) | 94.8% (94.1%, 95.4%) | 21.5% (19.9%, 23.2%) | 91.0% (90.1%, 91.8%) |
| Rotavirus | 32.1% (29.9%, 34.3%) | 77.0% (75.8%, 78.2%) | 21.4% (19.8%, 23.1%) | 89.1% (88.2%, 90.0%) | 20.4% (18.8%, 22.1%) | 90.3% (89.4%, 91.1%) |
| Pathogen Quantities | 66.7% (64.3%, 68.8%) | 91.4% (90.6%, 92.2%) | 40.3% (38.4%, 42.3%) | 81.2% (80.0%, 82.3%) | 42.6% (40.6%, 44.6%) | 80.3% (79.1%, 81.5%) |
| Symptoms | 72.1% (70.0%, 74.2%) | 89.7% (88.8%, 90.5%) | 27.2% (25.4%, 29.0%) | 88.1% (87.1%, 89.0%) | 40.6% (38.6%, 42.7%) | 76.9% (75.6%, 78.1%) |
| Pathogen + Symptoms | 75.9% (73.8%, 77.9%) | 91.9% (91.1%, 92.6%) | 44.7% (42.8%, 46.7%) | 84.5% (83.3%, 85.5%) | 54.4% (52.4%, 56.5%) | 80.7% (79.5%, 81.8%) |
| Host | 31.2% (29.1%, 33.5%) | 86.9% (85.9%, 87.8%) | 74.4% (72.7%, 76.1%) | 78.0% (76.7%, 79.2%) | 55.0% (53.0%, 57.1%) | 81.7% (80.6%, 82.8%) |
| Host + Symptoms | 43.9% (41.6%, 46.3%) | 86.8% (85.9%, 87.8%) | 77.8% (76.0%, 79.3%) | 80.8% (79.6%, 82.0%) | 67.1% (65.1%, 69.0%) | 80.5% (79.3%, 81.7%) |

### ii. Day 90 Re-hospitalization or Death Outcome

For the day 90 re-hospitalization or death outcome, on average 38.0% (95% CI: 36.9%, 39.2%) of children were recommended azithromycin treatment under the comprehensive rule with a threshold of a 2% absolute risk reduction (Table S4). Azithromycin was associated with a 2.4% lower risk of day 90 re-hospitalization or death (RD: -0.024 [95% CI: -0.041, -0.006]) among those for whom treatment was recommended compared to a 1.1% lower risk (RD: -0.011 [95% CI: -0.023, 0.000]) among those for whom treatment was not recommended. The characteristics of children recommended for treatment based on the day 90 re-hospitalization or death outcome differed from those based on the day 3 diarrhea outcome (Table 1, Figure S1). Specifically, while children recommended for treatment were more likely to have a bacterial etiology of diarrhea, the differences in the prevalence of bacteria attributed diarrhea between children recommended and not recommended for treatment were smaller compared to those for the day 3 diarrhea outcome. In contrast, differences in markers of malnutrition between those recommended and not recommended treatment were more pronounced for day 90 re-hospitalization or death, and SES was a strong determinant of the treatment recommendation. While similar proportions of children within each wealth quintile were recommended for treatment considering day 3 diarrhea, treatment was heavily skewed towards children in the lowest wealth quintiles when considering hospitalization or death (Table 1). Among children with moderate wasting or stunting, anthropometric measures were similar between children recommended for treatment versus those not, while differences in these measures between those recommended for treatment versus not were more pronounced among the subset of children with some or severe dehydration (Table S6).

The average treatment benefit for 90-day re-hospitalization or death among those recommended treatment was similar for all rules except the rule based on rotavirus only (RD: - 0.003 [95% CI: -0.044, 0.038]) (Table S4). Similarly to the day 3 diarrhea outcome, the treatment benefit for day 90 re-hospitalization or death was greatest under the *Shigella* rule (RD, -0.030 [95% CI: -0.070, 0.010]), but the proportion treated was small (8.4% [95% CI: 7.7%, 9.2%]), and approximately 66% of children with *Shigella*-attributed diarrhea were treated. The overall average benefits among all children when treated or not treated according to the rules were largest for the host and host + symptoms rules (at approximately a 1% absolute risk reduction) and were null for the rules based on a single pathogen (Table S4).

Rules that included child-level characteristics (i.e., host rule, host + symptoms rule, comprehensive rule) were more discriminating between children with respect to treatment benefit than rules based on pathogens and/or symptoms in that they tended to produce larger differences in average treatment benefit between those recommended for treatment versus not recommended for treatment (difference in RD: -0.013 [95% CI: -0.035, 0.009], -0.015 [95% CI: - 0.035, 0.006], -0.013 [95% CI: -0.034, 0.009], respectively) (Figure 1B, Table S4). Although the treatment benefit difference between those recommended and not recommended for treatment was also larger for the *Shigella* rule (-0.014 [95% CI: -0.056, 0.027]), the estimate of the average treatment benefit among those recommended for treatment was imprecise. The average sensitivity of the host rules and host + symptoms rules to identify children recommended for treatment by the comprehensive rule were considerably higher than that for other rules (74.4% and 77.8%, respectively), though their specificities were lower than other rules (78.0% and 80.8%, respectively) (Table 2, Table S5). Generally, the child-level expected benefit from the host rule (CCC: 0.65 [95% CI: 0.63, 0.66]) and host + symptoms rule (CCC: 0.73 [95% CI: 0.72, 0.74]) were better correlated with the child-level expected benefit from the comprehensive rule than other rules (Figure 2B), and treatment assignments from the host rule better aligned with those from the comprehensive rule (Figure S2).

### iii. Linear Growth Outcome

For the change in linear growth outcome, an average of 37.6% (95% CI: 36.5%, 38.9%) of children were recommended azithromycin treatment under the comprehensive rule using a threshold of 0.06 z-scores (Table S4). Azithromycin was expected to improve LAZ by 0.05 (95% CI: 0.01, 0.10) z-scores compared to the placebo among children recommended for treatment. The differences in the characteristics between children who were recommended and not recommended treatment according to the comprehensive rule for the linear growth outcome were less pronounced compared to those for the other outcomes (Table 1, Figure S1). Children recommended for treatment under the comprehensive rule were only slightly more likely to have diarrhea attributed to bacteria and had similar anthropometric measures compared to children not recommended for treatment. In contrast to rules for the other outcomes, children recommended treatment had shorter illness duration and were less likely to be dehydrated (Table 1).

None of the rules were particularly successful at distinguishing between children who did and did not benefit from treatment with respect to linear growth. Average changes in growth among those recommended treatment under the *Shigella* rule (0.08 z-scores [95% CI: 0.00, 0.16]), rotavirus rule (0.07 z-scores [95% CI: -0.01, 0.14]), and pathogen quantities rule (0.06 z-scores [95% CI: 0.01, 0.11]) were higher than that under the comprehensive rule (Table S4), and these rules had the largest differences in average benefit between those recommended for treatment versus those not (0.03 z-scores [95% CI: -0.05, 0.11], 0.02 z-scores [95% CI: -0.07, 0.10], 0.02 z-scores [95% CI: -0.04, 0.07], respectively), but all estimates were imprecise and not statistically different (Figure 1C, Table S4). The remaining rules, including the comprehensive, resulted in little to no difference between the average change in LAZ among those recommended for treatment versus that among those not recommended treatment. Sensitivity for identifying children recommended treatment by the comprehensive rule was modest and highest for the host + symptoms rule (67.1%), host rule (55.0%), and pathogen + symptoms rule (54.4%) while specificity was highest for the *Shigella* rule (91.0%), rotavirus rule (90.3%), and host rule (81.7%) (Table 2, Table S5). There was no correlation between the expected benefit or treatment assignment between the comprehensive rule and other rules (Figure 2C, Figure S2).

### iv. Threshold Comparison

We set thresholds at a 7% reduction in risk for day 3 diarrhea, 2% reduction in risk for day 90 re-hospitalization or death, and 0.06 difference in LAZ. In a sensitivity analysis, we examined how varying the threshold of clinical benefit that defines which children are recommended treatment impacted the proportion treated, expected benefits, and utility of different types of characteristics to define the rules. The proportion of children recommended for treatment by the comprehensive rule was dependent on the threshold of clinical benefit used to define which children were recommended treatment by the rule. For example, with a threshold of a 2% reduction in risk of diarrhea on day 3, the rule recommended approximately 85% of children for treatment versus recommending approximately 10% for treatment when the rule had a threshold of a 10% reduction in risk of diarrhea on day 3 (Figure 3). This pattern, in which the proportion of children recommended for treatment decreased as the threshold of clinical benefit increased, was consistent for each of the outcomes. The average benefit among children recommended for treatment generally increased as the clinical benefit threshold increased for the day 3 diarrhea and day 90 re-hospitalization or death outcomes, though estimates at more the largest thresholds lacked meaningful precision. In contrast, the average benefit in the linear growth outcome was similar across thresholds, with wide confidence intervals at the largest thresholds. Additionally, we found that the correlations between individual level expected benefit for 1) the pathogen quantities and comprehensive rules and 2) the host and comprehensive rules remained consistent over a range of thresholds (Figure S3).

**Figure 3:**
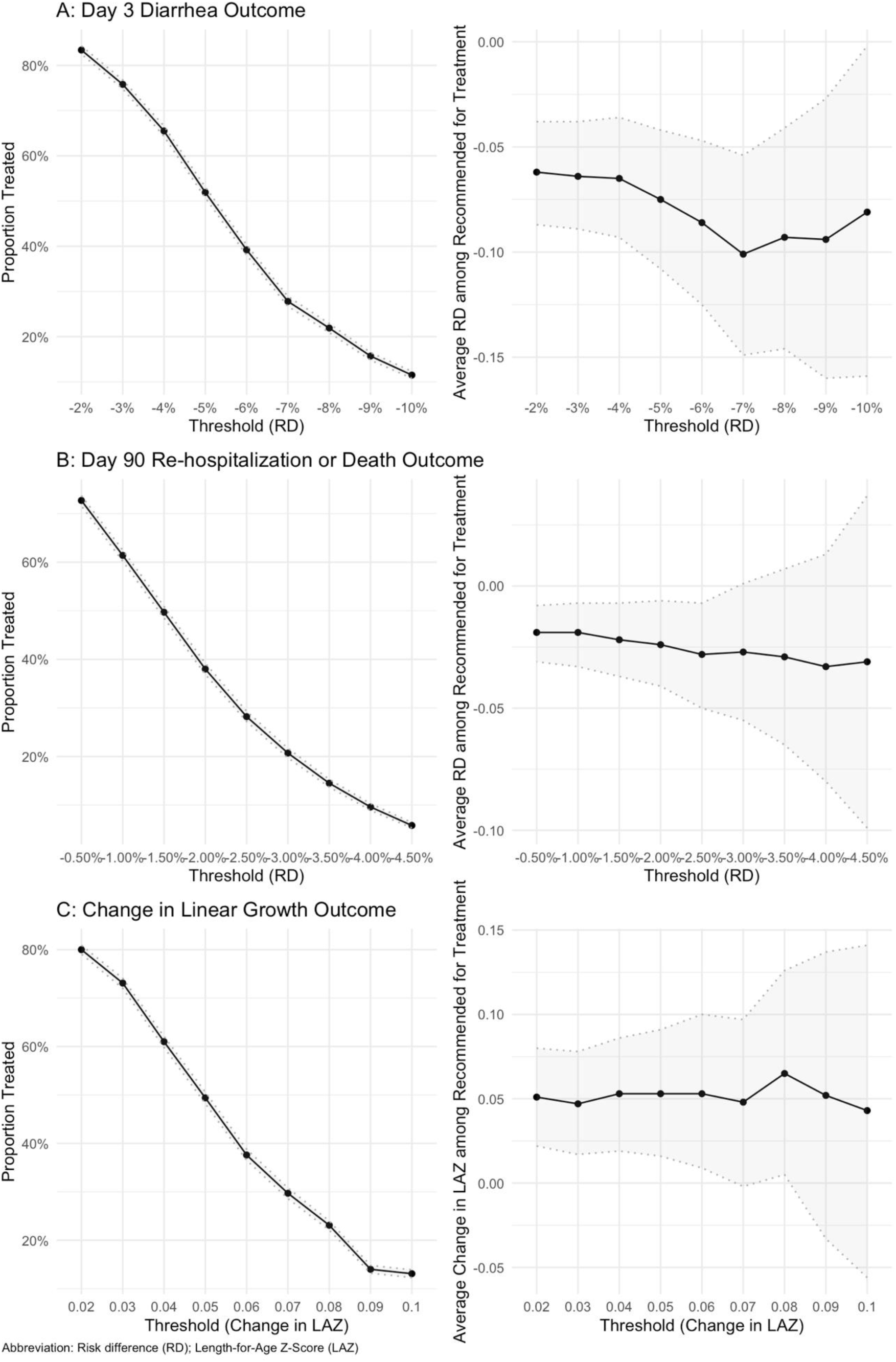
Proportion treated and average benefit (risk difference, RD) among those recommended for azithromycin under the comprehensive rule based on all known child and clinical characteristics with varying thresholds of clinical benefit to define the rule.

## Discussion

Although no overall survival benefit of azithromycin for moderately malnourished children with watery diarrhea was detected in the ABCD randomized trial (10), our secondary analysis demonstrated that children can be targeted for treatment based on their diagnostic, malnutrition, and/or sociodemographic characteristics to improve outcomes and limit antibiotic use. Personalized treatment rules, such as those evaluated here, may strike a better balance between expected benefit and risks of antibiotic resistance than both current WHO guidelines, which miss children likely to benefit from antibiotics, and current practice, in which the majority of watery diarrhea episodes are treated with antibiotics (8). While some of the rules included pathogen diagnostic data that are unlikely to be available at the point of care, other rules, such as the host rule or host + symptoms rule, only included information that would be easily observable in a clinical setting. While diagnostics were strong determinants of the effect of azithromycin on diarrhea duration, host characteristics were more relevant for identifying which children benefited most with respect to re-hospitalization or death. Therefore, appropriate targeting of antibiotics for children who would be expected to benefit in terms of the most severe outcomes may be achievable without widespread access to diagnostics. Deciding on the most appropriate personalized treatment rule in a given setting should weigh performance of the algorithm against feasibility with respect to data availability and limits on antibiotic use. For example, increasing the clinical benefit thresholds that defined the rules led to fewer children being recommended an antibiotic, which may be preferred in settings with concerns about antibiotic resistance.

The finding that bacterial pathogen diagnostics were most informative for the azithromycin treatment response with respect to diarrhea duration is logical. Because antibiotics directly affect growth of bacteria that are the proximal cause of diarrhea symptoms, it is biologically plausible that pathogen quantities are the most useful information to predict improvements in short-term recovery from diarrheal symptoms. On the other hand, the results that targeting children for azithromycin based on host characteristics, particularly anthropometric measures, is more effective to improve day 90 re-hospitalization or death is consistent with WHO guidelines that recommend routine treatment of antibiotics for children with SAM (14). Although the exact mechanism is not completely understood, hypotheses include that azithromycin reduces inflammatory responses and enteropathy that disrupt nutrient absorption, in addition to treating bacterial infections (15). In one of our sensitivity analyses, we found that among a subset of children with stunting or moderate wasting, 57% of children were recommended for treatment by the comprehensive rule compared to 36% in the full study population, highlighting malnutrition as a driver of treatment decision for the day 90 re-hospitalization or death outcome (Table S6). For the change in LAZ outcome, we generally observed little to no heterogeneity in average treatment benefit across rules, suggesting that benefit for linear growth from azithromycin treatment is similar regardless of pathogen profile, symptoms, malnutrition indicators, and sociodemographic characteristics. Gut inflammation is one of several plausible reasons for impaired growth outcomes (16), and the anti-inflammatory properties of azithromycin may similarly reduce inflammation and improve growth in all children.

The symptoms rule, which included information on days of diarrhea duration prior to enrollment, dehydration status, vomiting, and number of solid and loose stools in the prior 24 hours, did not recommend treatment for children similarly to the comprehensive rule for any outcome. This suggests that symptoms alone, information most proximal to the illness and readily available upon seeking care, may not be sufficiently informative for the decision to recommend antibiotics for children with acute watery diarrhea in these setting. Previous efforts to use clinical characteristics to inform treatment decisions have similarly found that epidemiologic factors like age, season, and anthropometry were most predictive of diarrhea etiology (17,18).

The treatment rules we developed recommended treatment for children whose estimated expected benefit exceeded a threshold. However, in some instances when these rules were evaluated in the validation data, the average benefit of treatment was estimated to be smaller than this threshold. This is a feature of the cross-validation procedure used, which seeks to evaluate the generalization performance of the estimated treatment rules. Although these rules were designed with the intention of treating only children who would have an expected benefit larger than the threshold, the ultimate estimated benefit may vary due to random variability. We sought to minimize some of this variability by averaging results across five repeated 10-fold cross-validation procedures.

This analysis was subject to limitations. First, the proportion recommended for treatment and estimates of average benefit were sensitive to the threshold chosen for treatment recommendation. For example, when we further increased the threshold for clinical benefit for the day 3 diarrhea outcome, a smaller proportion of children were treated, and the average treatment benefit among those recommended for treatment was stronger. However, the threshold for assigning treatment is inherently arbitrary and difficult to determine for outcomes like day 3 diarrhea which are non-severe and may not justify treatment with antibiotics alone. Second, because we used machine learning algorithms to predict child-level expected benefits, we could not characterize exactly which and how individual variables contributed to each rule. Therefore, interpretation of associations between specific variables and treatment decisions was descriptive, and we do not report interpretable criteria for recommending treatment. Third, these results may not be generalizable to all children with watery diarrhea because the data were collected among vulnerable children accompanied by some or severe dehydration, moderate wasting, or severe stunting between 2-23 months of age. Children with severe acute malnutrition, who may be at highest risk for severe outcomes of diarrhea, were excluded. Enrollment was also limited to seven study sites in Southeast Asia and Africa. While we intentionally excluded site from the rules to ensure they could be applied across settings, residual effects of site may be reflected in other characteristics such as SES and malnutrition indicators. Fourth, mortality benefit from azithromycin treatment was not explicitly measured because it was a particularly rare outcome (n=26 by day 90). Re-hospitalization and death by day 90 were combined as a single outcome to indicate severe outcomes while improving statistical power.

Using machine learning to compare alternative personalized treatment rules, we found that some diagnostic and host characteristics capture heterogeneity in azithromycin treatment response, and the most relevant characteristics differ by outcome. Our findings that pathogen quantities were important for treatment decisions related to day 3 diarrhea were expected. Interestingly, pathogen quantities were not necessary to identify which children to treat to prevent the most severe outcomes. Instead, measures of malnutrition and sociodemographic characteristics were more informative for the day 90 re-hospitalization or death outcome. These results highlight that diagnostics at the point of care in low resource settings may not be necessary to target treatment to prevent outcomes such as hospitalization or death but remain important for proximal outcomes such as illness duration. They also suggest antibiotic stewardship is feasible by demonstrating that antibiotics can be limited to the children who will benefit the most.

## Methods

### i. Study design

The ABCD trial enrolled 8,268 dehydrated or undernourished children with acute watery diarrhea across seven sites in Bangladesh, India, Kenya, Malawi, Mali, Pakistan, and Tanzania. Study procedures and methods for the ABCD trial have been described in prior publications (6,10). Briefly, children 2-23 months presenting to healthcare with acute watery diarrhea were eligible if they had some or severe dehydration, moderate wasting, or severe stunting and were excluded if they had dysentery, suspected cholera, severe acute malnutrition, or other infections requiring antibiotics. At enrollment, research staff collected clinical history (vomiting, number of loose stools in prior 24 hours, number of solid stools in prior 24 hours, number of days of diarrheal illness, dehydration status), measures of malnutrition (average middle-upper arm circumference, weight-for-age z-score, length-for-age z-score, and weight-for-length z-score), relevant demographic information (age, sex, socioeconomic status categorized in quintiles (based on country-specific wealth distributions in the Demographic and Health Survey (10)), number of children <5 in household), and fecal samples from all participants. Biospecimens, including whole stools and rectal swabs, for the first approximately 1,000 children enrolled at each site underwent pathogen testing by qPCR using the TaqMan Array Card platform. Details of the laboratory methods and quality control have been previously described (6). Our analysis considers both the presence and relative quantity (based on log-10 transformed qPCR cycle threshold values) of the 12 most-detected pathogens in the study: rotavirus, norovirus, adenovirus, astrovirus, sapovirus, ST ETEC, S*higella*, *Campylobacter*, tEPEC, *V. Cholerae*, *Salmonella*, and *Cryptosporidium*.

After enrollment, children were randomized 1:1 to receive a 3-day course of azithromycin (10 mg/kg) or placebo and were subsequently followed for up to 180 days. Follow-up visits occurred on days 2, 3, 45, 90, and 180 post-enrollment, during which vital status, anthropometric, clinical, and autopsy data were collected when relevant. This analysis focuses on 3 outcomes: 1) presence of diarrhea on day 3 following enrollment, 2) hospitalization following the enrollment-associated episode or death by day 90 following enrollment, and 3) change in LAZ between days 0 and 90.

### ii. Treatment Rules

We developed personalized treatment rules based on all available child and clinical characteristics as well as seven pre-specified sets of covariates based on the type of data and considering plausible scenarios of data availability and/or feasibility of implementation in real-world settings (Table 3). Comparison of results across rules allowed us to interrogate the relevance of different types of data as determinants of the heterogeneity in the treatment effects. The rule based on all available child, clinical, and diagnostic characteristics was considered the comprehensive rule since it included the most data compared to other rules.

**Table 3:** Description and rationale of personalized treatment rules.

| Treatment Rule | Variables that Determine Rule | Rationale |
| --- | --- | --- |
| **Comprehensive rule** | Rotavirus, norovirus, adenovirus, astrovirus, sapovirus, ST ETEC, *Shigella*, *Campylobacter*, tEPEC, *V. Cholerae*, *Salmonella*, and *Cryptosporidium* quantity and detection, vomit prior to enrollment, # loose stools in 24 prior to enrollment, # solid stools in 24 prior to enrollment, diarrhea duration prior to enrollment, dehydration status at enrollment, average middle-upper arm circumference, weight-for-age z-score, weight-for-length z-score, weight-for-age z-score, sex, age, SES quintile, # children <5 years in household | Uses all available data and is considered the best-informed rule |
| ***Shigella* rule** | *Shigella* quantity and detection | Approximates availability of a diagnostic for *Shigella* |
| **Rotavirus rule** | Rotavirus quantity and detection | Approximates availability of a diagnostic for rotavirus |
| **Pathogen quantities rule** | Rotavirus, norovirus, adenovirus, astrovirus, sapovirus, ST ETEC, *Shigella*, *Campylobacter*, tEPEC, *V. Cholerae*, *Salmonella*, and *Cryptosporidium* quantity and detection | Assumes diarrhea etiology alone determines the azithromycin benefit |
| **Symptoms rule** | Vomit prior to enrollment, # loose stools in 24 prior to enrollment, # solid stools in 24 prior to enrollment, diarrhea duration prior to enrollment, dehydration status at enrollment | Assumes clinical symptoms alone determine the azithromycin benefit |
| **Pathogen + symptoms rule** | Rotavirus, norovirus, adenovirus, astrovirus, sapovirus, ST ETEC, *Shigella*, *Campylobacter*, tEPEC, *V. Cholerae*, *Salmonella*, and *Cryptosporidium* quantity and detection, vomit prior to enrollment, # loose stools in 24 prior to enrollment, # solid stools in 24 prior to enrollment, diarrhea duration prior to enrollment, dehydration status at enrollment | Assumes host characteristics do not affect the azithromycin benefit |
| **Host rule** | Average middle-upper arm circumference, weight-for-age z-score, weight-for-length z-score, weight-for-age z-score, sex, age, SES quintile, # children <5 years in household | Assumes only host characteristics determine the azithromycin benefit |
| **Host + symptoms rule** | Vomit prior to enrollment, # loose stools in 24 prior to enrollment, # solid stools in 24 prior to enrollment, diarrhea duration prior to enrollment, dehydration status at enrollment, average middle-upper arm circumference, weight-for-age z-score, weight-for-length z-score, weight-for-age z-score, sex, age, SES quintile, # children <5 years in household | Evaluates all data that would be readily observable in clinical settings |

Treatment rules were developed using a robust ensemble machine learning-based procedure to estimate the expected benefit of azithromycin (i.e., additive risk reduction) to a child with respect to a particular outcome (e.g., presence of diarrhea on day 3 following enrollment) given a set of one or more characteristics of the child. Rules for treating children were defined based on this child-level expected benefit, whereby all children whose expected benefit exceeded a certain clinically relevant threshold were recommended azithromycin by the rule. For additional details on the machine learning pipeline for learning and evaluating treatment rules, see Supplementary Materials Methods.

### iii. Statistical Analysis

The study cohort was characterized by providing summary statistics of demographic and clinical characteristics. The primary analysis involved a two-step process involving randomly subsetting data into a training and validation sample. In the first step, we used the training sample to learn the child-level expected benefit for treatment given a set of input characteristics and developed a rule for treating children based on the level of expected benefit. In the second step, the validation sample was used to identify the children that would be recommended treatment given their expected benefit and to calculate the diagnostic performance of the rule.

For the day 3 diarrhea outcome, we focus our presentation on treatment rules that treat all children who have an expected benefit of at least 7% absolute reduction in risk of day 3 diarrhea, while treating no children with expected benefit ≤7% risk reduction. For the day 90 re-hospitalization or death outcome, we focus on treatment rules that treat children who have an expected 2% or greater absolute reduction in risk of hospitalization or death. For the change in LAZ outcome, we focus on rules that treat children who have an expected improvement of at least 0.06 units of LAZ. These outcome-specific cut-offs were based on a clinically relevant expected benefit and such that the proportion of children recommended treatment (30-45%) was likely to be acceptable considering concerns of antibiotic overuse and risks of antibiotic resistance. In sensitivity analyses, we varied these thresholds.

We quantified the impact of each treatment rule by estimating: (i) the proportion of children treated under the rule; (ii) the average benefit of treatment among those recommended treatment under the rule; (iii) the average benefit of treatment among those not recommended treatment under the rule; and (iv) the average benefit among all children of being treated according to the rule. While (ii) and (iii) compare the impact of azithromycin versus placebo among a subset of children, (iv) compares among all children the impact of treating all children according to the treatment rule (whereby some children are treated with azithromycin and others with placebo) versus treating all children with placebo. Estimate (iv) reflects comparing a scenario where the treatment algorithm was adopted for all children with watery diarrhea to a scenario where no child was treated. We then evaluate how well the rules discriminate between children in terms of their expected benefit by estimating the difference between the average benefit among those recommended for treatment (ii) with that among those not recommended for treatment (iii).

To avoid overfitting and to obtain an unbiased evaluation of the performance of our treatment rules, we utilized ten-fold cross-validation wherein data were randomly divided into ten distinct partitions, treatment rules were learned in a training sample and evaluated in a held-out validation sample. To reduce the randomness inherent in this sample splitting procedure (19), we then repeated the entire process five times and present point estimates and 95% confidence intervals for the average performance of the rules over the five repeated cross-validation procedures.

To better understand which types of data were strong determinants of the machine learned treatment rules, we present summary measures of child-level characteristics stratified by the decision to recommend treatment under the comprehensive rule. We also evaluate how well the rules approximate the comprehensive rule by estimating the sensitivity and specificity of each rule to identify children recommended for treatment by the comprehensive rule and the concordance correlation coefficient (CCC) of child-level expected benefit between the comprehensive and other rules.

To further interrogate the role of the threshold for our results, we estimated the CCC between individual level expected benefit for 1) the pathogen quantities and comprehensive rules and 2) the host and comprehensive rules across a range of thresholds. We assessed the sensitivity of the treatment rules to the threshold using the day 90 re-hospitalization and death outcome, which would be most relevant for treatment guideline changes.

## Data Availability Statement

The de-identified data that support the findings of this study are available from the corresponding author [S.S.K.] upon reasonable request and signed data access agreement upon publication.

## Acknowledgements

The ABCD trial and nested molecular diagnostics study were funded by the Bill & Melinda Gates Foundation (grant numbers OPP 1126331 and OPP 1179069). This study was funded by the Bill & Melinda Gates Foundation (INV-044317 to ETRM) and NIH/NIAID (R01AI185140 to ETRM).

## Author Contributions

E.T.R.M., D.B., J.A.P, and P.P. conceptualized and designed the study. S.S.K., A.C., E.T.R.M., and D.B. wrote the manuscript. A.C. and D.B. developed statistical methods. S.S.K. and A.C. performed data analyses. K.M., C.S., C.P.D., Q.D., N.B., K.K., S.O.S., S.S., B.O.S., J.W., F.Q., T.A., and A.D.C. supervised data acquisition. E.T.R.M. and D.B. supervised the study. All authors contributed to the revisions, interpretation of results, and completion of the final manuscript. All authors have read and approved the manuscript.

