## Supplementary Materials for "Personalized azithromycin treatment rules for children with watery diarrhea using machine learning"

Affiliations:

**Supplementary Methods**

To develop personalized treatment rules, we apply a machine learning approach to estimate the conditional average treatment effect (CATE) for a given set of covariates. For ease of interpretation, the CATE is referred to as the child-level expected benefit in the main manuscript. For example, to develop a comprehensive rule, we estimate the CATE given all covariate information available for a child. The CATE is a function of these covariates. When given a child’s covariate values, the CATE function returns the expected benefit (e.g., the average reduction in risk of day 3 diarrhea) for the child. We then threshold this expected benefit to define a treatment rule. That is, given a child’s covariate values, our rules recommend treatment for the child if the CATE-predicted expected benefit exceeds the threshold and recommends no treatment otherwise.

To estimate the CATE, we used a doubly-robust (DR) learner (1). In this approach a CATE estimate is built in two main steps: (i) estimation of nuisance regressions; (ii) estimation of a pseudo-outcome regression. In the first step, three separate regressions must be estimated. The first is an estimate of the conditional mean of the outcome of interest given azithromycin treatment assignment and all measured covariates. This model includes all covariates that are predictive or prognostic of the endpoint of interest. The second nuisance regression is the conditional probability of receiving azithromycin given this same set of covariates. Because the ABCD study was a 1:1 randomized trial, this probability (50%) is known by design and thus simple estimators may be used for the second nuisance regression. The third and final nuisance regression is the probability of having a non-missing outcome given azithromycin treatment assignment and all measured covariates.

In the second step of the DR learner, a pseudo-outcome regression is fit. Predictions from the nuisance regressions estimated in the first step are combined into *pseudo-outcomes* for each child according to the formula provided in Kennedy (2023) (1). This pseudo-outcome is then regressed against the (subset of) covariates with which we would like to build a treatment rule to generate an estimate of the CATE. This estimate of the CATE can then be thresholded, as described above, to generate a treatment rule.

The form the pseudo-outcome takes in the DR-learner is doubly robust, meaning that it is possible to incorrectly estimate either the conditional mean of the outcome OR the conditional probability of having a non-missing outcome, while still yielding a valid estimator of the pseudo-outcome regression in the second stage. This property allows our method to have increased flexibility and robustness in model estimation compared to other CATE estimation methods.

To further increase the flexibility of our method, we fit each regression model in the DR-Learner procedure using Super Learning. Super Learning employs cross-validation to combine multiple user-specified candidate regression models into an optimally weighted *ensemble* model (2). The resulting ensemble model is guaranteed to match or surpass accuracy of its individual candidate models (2). This approach allows us to test a wide range of models for estimating each nuisance regression and the pseudo-outcome regression (see supplementary tables S1-S3 and 3 in the main text for list of models used for each regression), while using a data-driven approach to select between the models. Super Learning has been shown to be an appropriate method for treatment rule estimation and uncover treatment effect heterogeneity more effectively than traditional approaches (3,4).

i. Characterizing the Performance of Rules

Given a treatment rule learned by thresholding a DR-learner of the CATE, we evaluated the performance of the rule on several axes: the proportion of children recommended treatment under the rule, the average effect (e.g., additive risk reduction) of azithromycin treatment in the subpopulation recommended for treatment, the average effect of azithromycin treatment in the subpopulation *not* *recommended* for treatment, and the additive effect of treating all children according to the rule vs. treating no children. We used sample proportions to estimate the proportion of children treated by the rule. All other effect-related performance metrics were estimated using cross-validated Augmented Inverse Probability of Treatment Weighted (AIPTW) estimators, also known as doubly-debiased machine learning estimators of the effects (5). Similarly as with the DR-learner of the CATE, such estimators enjoy a double-robustness property. The effect estimators are built using the same nuisance regression estimates as are used to construct the DR-learner and are constructed such that it is possible to incorrectly estimate either the conditional mean of the outcome OR the conditional probability of having a non-missing outcome, while still yielding a valid estimator of the effect of interest.

ii. Cross-Validation for Rule Development and Evaluation

To guard against the possibility of machine learning over-fitting, we utilize extensive cross-validation to separate the process of (i) learning the treatment rules and (ii) estimating their performance. Further, in line with the recommendations of Kennedy (2023) (1), while learning the treatment rule, we separate the process of (i.a) estimating the nuisance regressions and (i.b) estimating the pseudo-outcome regression.

For the purposes of separating (i) from (ii), we utilized 10-fold cross-validation. That is, we divided the data into ten mutually exclusive partitions. We utilized nine-tenths of the data to learn a treatment rule and then used the remaining 1/10 to estimate its performance. We then repeated this process holding out each of the ten partitions in turn. Performance of rules was then averaged over the ten folds.

For the purposes of separating (i.a) from (i.b), we utilized a nested layer of 3-fold cross-validation. That is, given 9/10 of the data, we partition these data further into 3 mutually exclusive sub-partitions. Two of these sub-partitions are used to estimate nuisance regressions, while the remaining partition is used to construct pseudo-outcomes and fit the pseudo-outcome regression. Finally, the treatment rule learned using the 9/10 of data is defined by thresholding the average CATE estimate from each of the 3 inner sub-partitions.

The method implementation used to run all analysis can be found in the *drotr* R package on GitHub (6).

**Overview of covariates and notation**

**Treatment (A):**

Azithromycin vs. placebo

**Outcome (Y):**

1. Diarrhea on day 3 of trial
2. Re-hospitalization or death by day 90 of trial
3. Length for age z-score at day 90 of trial

**Covariates (W):**

1. Pathogen quantity (quantity and any detection)
a. Rotavirus
b. Norovirus
c. Adenovirus 40/41
d. Sapovirus
e. Astrovirus
f. ST-ETEC
g. *Shigella*
h. *Campylobacter jejuni/coli*
i. Typical EPEC
j. *Vibrio Cholera*
k. *Salmonella*
l. *Cryptosporidium*
2. Illness characteristics
   a. Vomiting prior to enrollment
   b. Number of loose stools in the 24 hours prior to enrollment
   c. Number of solid stools in the 24 hours prior to enrollment
   d. Duration (days) of diarrhea illness prior to enrollment
   e. Dehydration status at enrollment
3. Malnutrition indicators at enrollment
   a. Middle-upper arm circumference
   b. Weight for age z-score
   c. Length for age z-score
   d. Weight for length z-score
4. Sociodemographic characteristics
   a. Site
   b. Sex
   c. Age in months
   d. SES Quintile
   e. Number of children <5 in household
5. Other
   a. Month of the year enrolled
   b. Rotavirus season
      i. Bangladesh: November – February (7)
      ii. Kenya: January – March and June – September (8)
      iii. Malawi: May – October (9)
      iv. Mali: March – June (10)
      v. India: December – February (11)
      vi. Tanzania: May – October (12)
      vii. Pakistan: November – March (13)

**Supplementary Table 1:** Built-in and manually defined models used to fit outcome model with SuperLearner with all covariates W^1^

**Description**

Generalized linear model

Lasso

Lasso with two-way interactions

Gradient boosting

Random forest

Multivariate adaptive regression splines

Generalized linear model with interactions between treatment and site and treatment and *Shigella* pathogen quantity

Generalized linear model with interactions between treatment and site and treatment and rotavirus pathogen quantity

Generalized linear model with interactions between treatment and site and treatment and 2 pathogen quantities (*Shigella*, rotavirus)

Generalized linear model with interactions between treatment and site and treatment and rotavirus season

Generalized linear model with interactions between treatment and site and treatment and month enrolled

Generalized linear model with interactions between treatment and site and treatment and top 12 pathogen quantities

Generalized linear model with interactions between treatment and site and treatment and vomiting

Generalized linear model with interactions between treatment and site, treatment and top 12 pathogen quantities, and treatment and illness characteristics

Generalized linear model with interactions between treatment and site, treatment and malnutrition indicators, and treatment and sociodemographic characteristics

Generalized linear model with interactions between treatment and site, treatment and illness characteristics, treatment and malnutrition indicators, and treatment sociodemographic characteristics

Generalized linear model with interactions between treatment and all covariates W^1^

^1^W contains all variables listed in “Covariates (W)” section

**Supplementary Table 2:** Built-in and manually defined models used to fit treatment model with SuperLearner

**Description**

Mean

Logistic regression with all covariates (W^1^)

^1^W contains all variables listed in “Covariates (W)” section

**Supplementary Table 3:** Built-in and manually defined models used to fit missingness model with SuperLearner

| **Description** | **Justification** |
| --- | --- |
| Mean | Primary reason for outcome missingness was timing of IRB approval by study site |
| Logistic regression with site and month |  |
| Logistic regression with interaction between site and month |  |

**Supplemental Table 4**: Proportion of watery diarrhea episodes treated with azithromycin and average benefits of personalized treatment rules for day 3 diarrhea, day 90 re-hospitalization or death, and change in linear growth averaged across 5 seeds.

|  | **Proportion Treated**  **(95% CI)** | | **Average Benefit among Recommended for Treatment (95% CI)** | | **Average Benefit among Not Recommended for Treatment**  **(95% CI)** | | **Difference in Average Benefit Among Recommended vs Not (95% CI)** | **Average Benefit of Treatment Rule (95% CI)** |
| --- | --- | --- | --- | --- | --- | --- | --- | --- |
| **Day 3 Diarrhea** | |  | |  | |  | | |
| Comprehensive | 27.8% (26.7%, 28.9%) | | -0.101 (-0.149, -0.054) | | -0.037 (-0.063, -0.011) | | -0.065 (-0.119, -0.011) | -0.026 (-0.039, -0.013) |
| Shigella | 15.9% (15.0%, 16.8%) | | -0.135 (-0.202, -0.068) | | -0.040 (-0.064, -0.016) | | -0.095 (-0.166, -0.024) | -0.020 (-0.030, -0.009) |
| Rotavirus | 32.7% (32.0%, 33.4%) | | -0.038 (-0.142, 0.066) | | -0.036 (-0.084, 0.012) | | -0.002 (-0.116, 0.113) | -0.013 (-0.026, 0.000) |
| Pathogen Quantities | 25.3% (24.3%, 26.4%) | | -0.094 (-0.140, -0.047) | | -0.042 (-0.068, -0.015) | | -0.052 (-0.106, 0.002) | -0.021 (-0.032, -0.010) |
| Symptoms | 18.7% (17.8%, 19.6%) | | -0.089 (-0.159, -0.019) | | -0.049 (-0.073, -0.025) | | -0.040 (-0.114, 0.034) | -0.012 (-0.024, 0.000) |
| Pathogen + Symptoms | 27.3% (26.2%, 28.3%) | | -0.095 (-0.143, -0.047) | | -0.039 (-0.065, -0.012) | | -0.056 (-0.111, -0.001) | -0.024 (-0.036, -0.011) |
| Host | 23.8% (22.8%, 24.8%) | | -0.072 (-0.130, -0.014) | | -0.051 (-0.076, -0.026) | | -0.021 (-0.084, 0.043) | -0.013 (-0.026, -0.001) |
| Host + Symptoms | 25.8% (24.8%, 26.9%) | | -0.055 (-0.108, -0.001) | | -0.054 (-0.079, -0.030) | | 0.00 (-0.060, 0.058) | -0.013 (-0.026, 0.001) |
| **Day 90 Re-hospitalization/Death** | | |  | |  | |  |  |
| Comprehensive | 38.0% (36.9%, 39.2%) | | -0.024 (-0.041, -0.006) | | -0.011 (-0.023, 0.000) | | -0.013 (-0.034, 0.009) | -0.008 (-0.015, -0.002) |
| Shigella | 8.4% (7.7%, 9.2%) | | -0.030 (-0.070, 0.010) | | -0.016 (-0.026, -0.005) | | -0.014 (-0.056, 0.027) | -0.002 (-0.006, 0.002) |
| Rotavirus | 13.6% (12.8%, 14.4%) | | -0.003 (-0.044, 0.038) | | -0.018 (-0.028, -0.007) | | 0.015 (-0.028, 0.057) | -0.001 (-0.004, 0.003) |
| Pathogen Quantities | 30.5% (29.5%, 31.6%) | | -0.023 (-0.043, -0.003) | | -0.014 (-0.025, -0.002) | | -0.009 (-0.033, 0.014) | -0.006 (-0.012, 0.000) |
| Symptoms | 24.1% (23.1%, 25.1%) | | -0.020 (-0.050, 0.009) | | -0.016 (-0.027, -0.006) | | -0.004 (-0.035, 0.027) | -0.003 (-0.009, 0.003) |
| Pathogen + Symptoms | 30.7% (29.6%, 31.8%) | | -0.015 (-0.037, 0.006) | | -0.015 (-0.026, -0.004) | | 0.000 (-0.024, 0.024) | -0.005 (-0.011, 0.002) |
| Host | 41.5% (40.4%, 42.7%) | | -0.022 (-0.040, -0.004) | | -0.010 (-0.022, 0.003) | | -0.013 (-0.035, 0.009) | -0.010 (-0.017, -0.003) |
| Host + Symptoms | 41.7% (40.5%, 42.8%) | | -0.025 (-0.042, -0.008) | | -0.010 (-0.022, 0.002) | | -0.015 (-0.035, 0.006) | -0.010 (-0.017, -0.003) |
| **Change in Linear Growth** |  | |  | |  | |  |  |
| Comprehensive | 37.6% (36.5%, 38.9%) | | 0.05 (0.01, 0.10) | | 0.05 (0.02, 0.09) | | 0.00 (-0.06, 0.06) | 0.02 (0.00, 0.03) |
| Shigella | 12.0% (11.2%, 12.8%) | | 0.08 (0.00, 0.16) | | 0.05 (0.02, 0.08) | | 0.03 (-0.05, 0.11) | 0.01 (0.00, 0.02) |
| Rotavirus | 13.0% (12.2%, 13.8%) | | 0.07 (-0.01, 0.14) | | 0.05 (0.02, 0.08) | | 0.02 (-0.07, 0.10) | 0.01 (0.00, 0.02) |
| Pathogen Quantities | 41.7% (30.6%, 32.8%) | | 0.06 (0.01, 0.11) | | 0.05 (0.01, 0.08) | | 0.02 (-0.04, 0.07) | 0.02 (0.00, 0.03) |
| Symptoms | 34.5% (33.4%, 35.6%) | | 0.05 (0.00, 0.10) | | 0.05 (0.02, 0.08) | | 0.00 (-0.06, 0.06) | 0.02 (0.00, 0.03) |
| Pathogen + Symptoms | 34.9% (33.8%, 36.0%) | | 0.05 (0.00, 0.09) | | 0.05 (0.02, 0.09) | | 0.00 (-0.06, 0.05) | 0.02 (0.00, 0.03) |
| Host | 35.8% (34.7%, 36.9%) | | 0.06 (0.00, 0.11) | | 0.06 (0.02, 0.09) | | 0.00 (-0.07, 0.06) | 0.01 (0.00, 0.03) |
| Host + Symptoms | 39.2% (38.1%, 40.4%) | | 0.05 (0.01, 0.10) | | 0.05 (0.02, 0.09) | | 0.00 (-0.05, 0.06) | 0.02 (0.00, 0.03) |

**Supplemental Table 5**: Overlap in treated individuals under comprehensive rule versus other rules

|  | **Day 3 Diarrhea** | | **Day 90 Re-hospitalization/Death** | | **Change in Linear Growth** | |
| --- | --- | --- | --- | --- | --- | --- |
|  | Comprehensive | | Comprehensive | | Comprehensive | |
|  | Treated | Untreated | Treated | Untreated | Treated | Untreated |
| **Shigella** |  |  |  |  |  |  |
| Treated | 904 | 275 | 361 | 222 | 488 | 398 |
| Untreated | 786 | 4,727 | 2,078 | 4,031 | 1,781 | 4,025 |
| **Rotavirus** |  |  |  |  |  |  |
| Treated | 542 | 1,149 | 523 | 463 | 462 | 429 |
| Untreated | 1,148 | 3,853 | 1,916 | 3,790 | 1,807 | 3,994 |
| **Pathogen Quantities** |  |  |  |  |  |  |
| Treated | 1,125 | 430 | 943 | 800 | 966 | 870 |
| Untreated | 565 | 4,572 | 2,439 | 3,453 | 1,303 | 3,553 |
| **Symptoms** |  |  |  |  |  |  |
| Treated | 1,219 | 516 | 663 | 508 | 922 | 1,022 |
| Untreated | 471 | 4,486 | 1,776 | 3,745 | 1,347 | 3,401 |
| **Pathogen + Symptoms** |  |  |  |  |  |  |
| Treated | 1,283 | 407 | 1,091 | 661 | 1,235 | 855 |
| Untreated | 407 | 4,595 | 2,439 | 3,592 | 1,034 | 3,568 |
| **Host** |  |  |  |  |  |  |
| Treated | 528 | 655 | 1,815 | 936 | 1,249 | 808 |
| Untreated | 1,162 | 4,347 | 624 | 3,317 | 1,020 | 3,615 |
| **Host + Symptoms** |  |  |  |  |  |  |
| Treated | 742 | 658 | 1,895 | 816 | 1,522 | 861 |
| Untreated | 948 | 4,344 | 544 | 3,437 | 747 | 35,62 |

**Supplemental Table 6:** Descriptive characteristics by treatment recommendation under the comprehensive rule with respect to day 90 re-hospitalization or death among subsets of children with dehydration and stunting/wasting

|  | All children | | Among dehydrated | | Among stunted/moderately wasted | |
| --- | --- | --- | --- | --- | --- | --- |
|  | Not Recommended  (64%) | Recommended Treatment  (36%) | Not Recommended  (69%) | Recommended Treatment  (31%) | Not Recommended (43%) | Recommended Treatment  (57%) |
|  | N (Col %); Mean (SD) | N (Col %); Mean (SD) | N (Col %); Mean (SD) | N (Col %); Mean (SD) | N (Col %); Mean (SD) | N (Col %); Mean (SD) |
| Total | 4,253 | 2,439 | 2,511 | 1,128 | 1,652 | 2,208 |
| Bacteria Attributed Diarrhea |  |  |  |  |  |  |
| Likely | 994 (23%) | 900 (37%) | 642 (26%) | 374 (33%) | 389 (24%) | 747 (34%) |
| Possible | 787 (19%) | 366 (15%) | 398 (16%) | 159 (14%) | 359 (22%) | 348 (16%) |
| Unlikely | 2,472 (58%) | 1,173 (48%) | 1,471 (59%) | 595 (53%) | 904 (55%) | 1,113 (50%) |
| Likely *Shigella* Diarrhea | 345 (8%) | 500 (21%) | 253 (10%) | 157 (14%) | 132 (8%) | 407 (18%) |
| Likely ST ETEC Diarrhea | 521 (12%) | 368 (15%) | 312 (12%) | 173 (16%) | 194 (12%) | 319 (14%) |
| Likely tEPEC Diarrhea | 112 (3%) | 103 (4.2%) | 64 (2.5%) | 60 (5%) | 63 (4%) | 77 (4%) |
| Likely *V. Cholerae* Diarrhea | 97 (2.3%) | 77 (3.2%) | 84 (3%) | 43 (4%) | 18 (1%) | 57 (3%) |
| Likely *Campylobacter* Diarrhea | 4 (<0.1%) | 2 (<0.1%) | 2 (<0.1%) | 2 (0.2%) | 3 (0.2%) | 1 (<0.1%) |
| Likely *Salmonella* Diarrhea | 26 (1%) | 26 (1%) | 16 (1%) | 5 (0.4%) | 22 (1%) | 20 (1%) |
| Likely Rotavirus Diarrhea | 674 (16%) | 737 (30%) | 420 (17%) | 306 (27%) | 295 (18%) | 604 (27%) |
| Likely *Cryptosporidium* Diarrhea | 398 (9%) | 241 (10%) | 226 (9%) | 127 (11%) | 114 (9%) | 243 (11%) |
| Age (Months) | 11.60 (5.55) | 11.69 (4.73) | 11.46 (5.58) | 10.90 (4.74) | 11.80 (5.61) | 11.65 (4.89) |
| Female Sex | 1,937 (46%) | 1,151 (47%) | 1,139 (45%) | 523 (46%) | 720 (44%) | 1,082 (49%) |
| Study Site |  |  |  |  |  |  |
| Bangladesh | 376 (8.8%) | 622 (26%) | 114 (4.5%) | 135 (12%) | 279 (17%) | 563 (25%) |
| India | 608 (14%) | 390 (16%) | 258 (10%) | 225 (20%) | 328 (20%) | 400 (18%) |
| Kenya | 855 (20%) | 159 (6.5%) | 730 (29%) | 201 (18%) | 71 (4.3%) | 106 (4.8%) |
| Malawi | 462 (11%) | 229 (9.4%) | 365 (15%) | 172 (15%) | 107 (6.5%) | 145 (6.6%) |
| Mali | 632 (15%) | 368 (15%) | 157 (6.3%) | 74 (6.6%) | 448 (27%) | 421 (19%) |
| Pakistan | 498 (12%) | 497 (20%) | 169 (6.7%) | 115 (10%) | 325 (20%) | 491 (22%) |
| Tanzania | 822 (19%) | 174 (7.1%) | 718 (29%) | 206 (18%) | 94 (5.7%) | 82 (3.7%) |
| SES Quintile |  |  |  |  |  |  |
| First | 223 (5.2%) | 641 (26%) | 156 (6.2%) | 134 (12%) | 167 (10%) | 493 (22%) |
| Second | 798 (19%) | 294 (12%) | 374 (15%) | 91 (8.1%) | 349 (21%) | 410 (19%) |
| Third | 586 (14%) | 485 (20%) | 309 (12%) | 272 (24%) | 240 (15%) | 392 (18%) |
| Fourth | 972 (23%) | 807 (33%) | 608 (24%) | 470 (42%) | 324 (20%) | 597 (27%) |
| Fifth | 1,674 (39%) | 212 (8.7%) | 1,064 (42%) | 161 (14%) | 572 (35%) | 316 (14%) |
| # <5 Years in Household | 1.63 (0.92) | 1.86 (1.24) | 1.61 (0.95) | 1.71 (0.99) | 1.74 (1.07) | 1.86 (1.22) |
| Length for Age Z-Score | -1.33 (1.37) | -1.71 (1.29) | -0.94 (1.26) | -1.19 (1.18) | -2.01 (1.35) | -1.91 (1.26) |
| Weight for Age Z-Score | -1.44 (1.32) | -2.06 (1.01) | -0.77 (1.19) | -1.45 (1.10) | -2.52 (0.78) | -2.35 (0.69) |
| Weight for Length Z-Score | -0.91 (1.27) | -1.55 (1.02) | -0.31 (1.13) | -1.11 (1.07) | -1.85 (0.85) | -1.79 (0.92) |
| Middle-Upper Arm Circumference | 13.34 (1.24) | 12.66 (0.92) | 13.90 (1.17) | 13.17 (0.99) | 12.38 (0.82) | 12.41 (0.71) |
| # Loose Stools in 24 Hours | 6.69 (3.33) | 8.35 (4.43) | 6.69 (3.40) | 8.55 (4.72) | 7.15 (3.88) | 7.79 (3.91) |
| Illness Duration (Days) | 2.46 (1.95) | 2.60 (2.02) | 2.43 (1.94) | 2.30 (1.91) | 2.47 (1.92) | 2.79 (2.12) |
| Dehydration Status |  |  |  |  |  |  |
| None | 1,600 (38%) | 1,453 (60%) | 0 (0%) | 0 (0%) | 1,329 (80%) | 1,707 (77%) |
| Some | 2,398 (56%) | 887 (36%) | 2,261 (90%) | 1,024 (91%) | 213 (13%) | 463 (21%) |
| Severe | 255 (6.0%) | 99 (4.1%) | 250 (10.0%) | 104 (9.2%) | 110 (6.7%) | 38 (1.7%) |
| Vomit during Illness | 40 (0.9%) | 145 (5.9%) | 39 (1.6%) | 129 (11%) | 15 (0.9%) | 61 (2.8%) |

**Supplemental Figure 1**: Association between child-level expected benefit from comprehensive rule, stratified by treatment recommendation and select characteristics

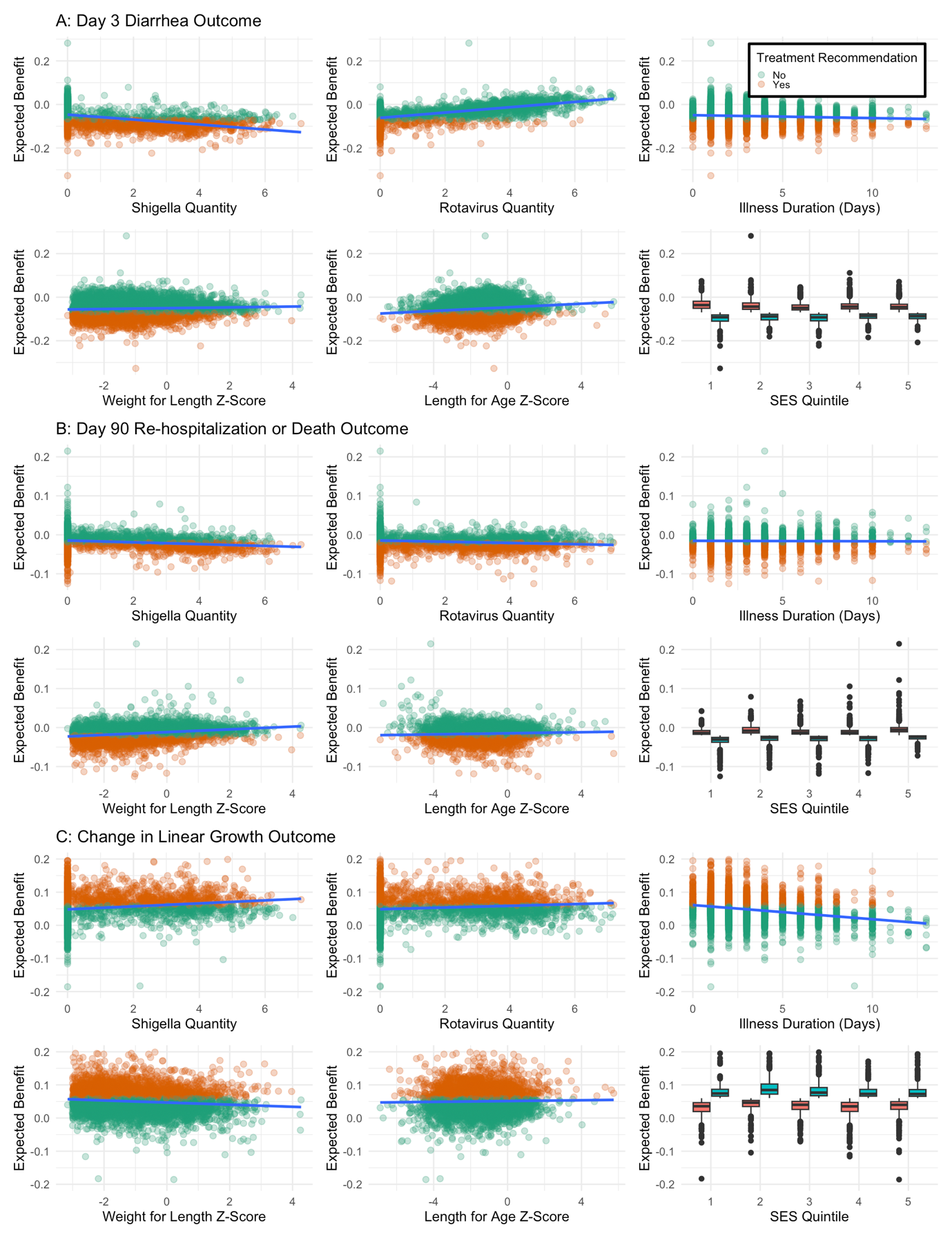

**Supplemental Figure 2**: Child-level expected benefit under comprehensive rule stratified by treatment recommendation under alternative rule

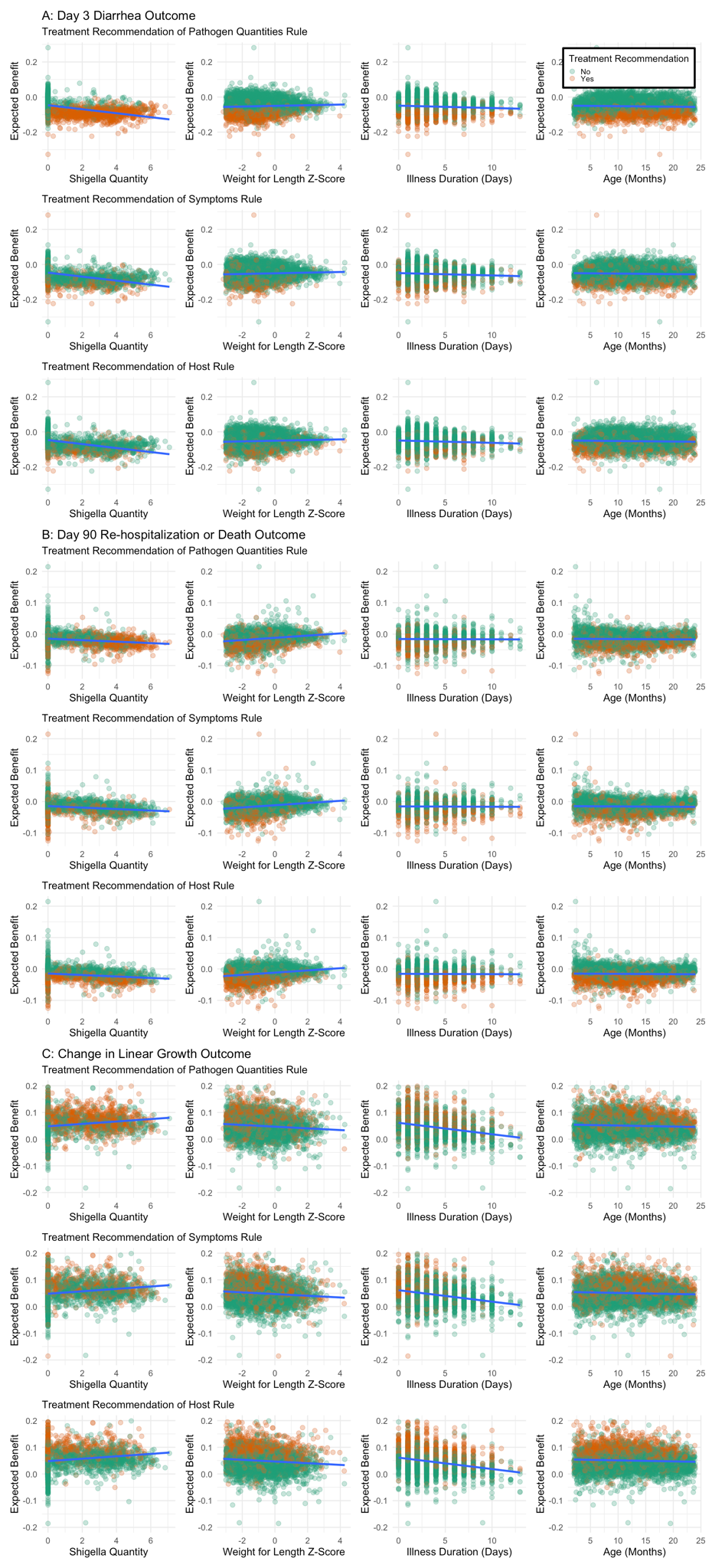

**Supplemental Figure 3:** Concordance correlation coefficient between child-level expected benefits from the 1) pathogen quantities vs comprehensive rule and 2) host vs comprehensive rule with respect to the day 90 re-hospitalization or death outcome across thresholds of clinical benefit

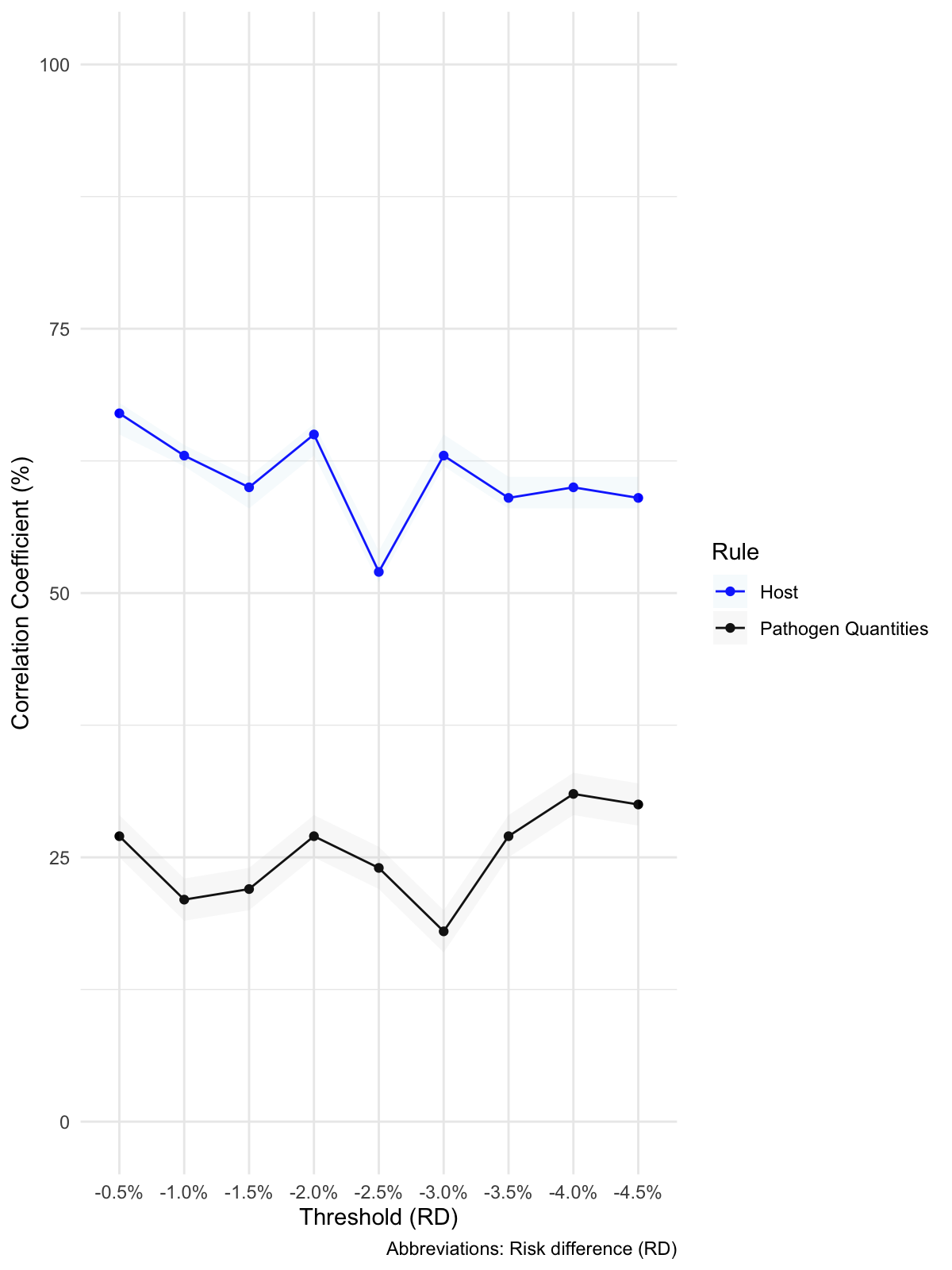
